# Study on the Correlation Between Biliary Tract and Intestinal Flora and the Formation of Gallstones

**DOI:** 10.1101/2022.06.28.22277035

**Authors:** Min Xie, Xue-ling Zhang, Yue Wu, Jia-huan Sun, Wei Yu, Pei-lin Cui

## Abstract

**Objective:** In recent years, the incidence of gallstones and their complications has increased, bringing a heavy burden to patients, emphasizing the need to explore the pathogenesis of gallstones. Evidences suggest that the formation of gallstones is closely related to the biliary tract and the gut flora. This study aims to reveal the diversity and abundance of intestinal flora in patients with biliary stones, investigate the relationship between the structure of gallstone formation and its flora, and preliminarily research gene function annotation and metabolic pathways.

**Methods:** The subjects were 21 eligible gallstone patients undergoing surgery and 20 eligible gallstone-free patients admitted to Beijing Tiantan Hospital, Capital Medical University, from November 2019 to November 2020. Gallstones (GSS group), bile (GSZ group), gallbladder mucosa (GSN group), feces (GSF group) samples were collected from the gallstone group, as well as feces from the control group (HF group). High-throughput sequencing of the V3-V4 regions of the 16S rRNA gene was performed by the Illumina HiSeq platform, bioinformatics analysis was performed on the sequencing results.

**Results:** 1. The age, body mass index (BMI) and indirect bilirubin (IBil) of gallstone patients were higher than gallstone-free patients (P < 0.05). 2. A total of 23 427 Operational Taxonomic Units (OTUs) were identified in this study, with a mean ± standard deviation of 340±93, including 4 095 from gallstones (GSS group), 3 065 from bile (GSZ group), 4 687 from gallbladder mucosa (GSN group), and 5 203 from feces (GSF group). 6 377 OTUs were identified from the feces of the gallstone-free control group (HF group). 3. There was no significant difference in the diversity and phylum composition of intestinal flora between gallstone patients and the control group (P > 0.05); however, at the genus level, Achromobacter (P=0.010), Faecalibacterium (P=0.042), Lachnospira (P=0.011) were significantly reduced, while Enterococcus (P=0.001) was significantly increased. 4. The diversity and composition of biliary flora (stone, bile, mucosa) among patients with gallstones have no statistical differences (P > 0.05). The diversity and composition between the biliary and intestinal microflora in gallstones patients have statistical differences: (1) The diversity of biliary flora was significantly higher than the intestinal flora (Simpson index, P < 0.05). (2) At the phylum level, the abundance of Proteobacteria in the bile duct (stone, bile and mucosa) was significantly higher, while Firmicutes and Bacteroidetes were significantly lower than in the intestinal tract (P < 0.05). (3) At the genus level, the abundance of Acinetobacter in the biliary tract was significantly higher, while Bacteroides, Faecalibacterium, Lachnoclostridium and Subdoligranulumbacteria were significantly lower than in the intestinal tract (P < 0.05). 5. The patient’s stone, bile and gallbladder mucosa shared more than 90% of OTUs. The shared OTUs of intestinal flora between gallstones patients and the control group was greater than 85%, while the five groups of samples shared more than 60% of OTUs. 6. LefSe showed that LDA > 4 in the biliary tract was Gammaproteobacteria, Pseudomonadales, Moraxellaceae, Acinetobacter, Betaproteobacteria, Burkholderiales and Prevotella that all belong to Proteobacteria.

**Conclusion:** The intestinal flora of patients with gallstones and without gallstones exhibited significant bacterial heterogeneity at the genus level. Compared with the intestinal flora of patients with gallstones, the biliary flora exhibited higher diversity. There were significant differences in the bacterial community structure at the phylum and genus levels. The biliary tract (stone, bile, mucosa) and intestinal flora of patients with gallstones have overlaps and differences, which provides the foothold for future studies on the biliary tract flora.

## Introduction

Gallstones are deposits of crystals in the biliary system, including the gallbladder or bile duct. Gallstones are very common worldwide, affecting 10-20% of the adult population^[1]^. With the change in people’s dietary structure and the acceleration of the aging process, the incidence of this disease is on the rise. However, surgery remains the mainstay of treatment. Therefore, more emphasis should be placed on new prevention strategies against the formation of gallstones.

The clinical manifestations of gallstones depend on the location and size of the calculi, with 5% of patients with gallstones developing acute cholecystitis, suppurative cholangitis, severe acute pancreatitis, biliary fistula and other serious complications^[2,3]^. In addition, gallstones have been associated with an increased risk of many chronic diseases, such as diabetes and cardiovascular disease^[4,5]^. More importantly, overwhelming evidence substantiates that the transformation from benign hyperplasia to malignant transformation of gallbladder mucosal epithelial cells caused by long-term gallstone stimulation may be one of the causes of gallbladder cancer^[6,7]^. Recent studies have shown that patients with asymptomatic gallstones have a significantly increased risk of right-sided colon cancer after 15 years^[8,9]^.

The current understanding of the pathogenesis of gallstones is very complex, mainly involving local and systemic factors. Local factors include gallbladder wall motility disorder, local persistent immune-mediated inflammation, mucin secretion and accumulation, cholesterol supersaturation, solid crystal precipitation, etc.^[10,11]^. On the other hand, systemic factors generally include gene polymorphism, epigenetic factors, expression and activity of nuclear receptors, insulin resistance, slow intestinal peristalsis, increased cholesterol absorption, etc.^[10,12]^. Some patients with gallstones suffer from discomfort such as belching, abdominal pain, abdominal distension and constipation for a long time. In recent years, much emphasis has been placed on understanding whether the formation of gallstones is related to intestinal flora disorders.

## Methods

The study subjects were 21 patients that underwent gallstone surgery at the Beijing Tiantan Hospital affiliated with Capital Medical University and 20 healthy patients without gallstones who underwent physical examination from November 2019 to November 2020. Gallstone (GSS group), bile (GSZ group), gallbladder mucosa (GSN group) and feces (GSF) specimens were collected from the gallstone group, and feces (HF group) were collected from the control group. This study were approved by the institutional research ethics committee of Beijing Tiantan Hospital, Capital Medical University.

### 1.1 Inclusion Criteria

Patients with gallstones were confirmed by abdominal B-ultrasound or CT examination.

### 1.2 Exclusion Criteria

1) Patients with severe bacteremia and septicemia received antibiotics within the last 3 months; 2) Presence of other serious metabolic diseases (such as severe obesity, uncontrollable hyperlipidemia, diabetes); 3) Patients that took a large dose of probiotics 3 months before the study; 4) Patients that took somatostatin or other drugs affecting gallstone formation; 5) Pregnant women or long-term contraceptive users; 6) Patients who underwent endoscopic retrograde cholangiopancreatography (ERCP) and intestinal surgery.

### 2. Sample Collection

#### 2.1 Gallstone group

##### 2.1.3 Gallstones

After the gallstones were collected during surgery, the surface of the stones was cleaned with sterile normal saline, and the specimens were stored in a sterile cryopreservation tube in liquid nitrogen at -80°C.

##### 2.1.2 Bile collection and treatment

after obtaining access to the abdominal cavity, the gallbladder was exposed, the bottom of the gallbladder was punctured with a sterile syringe, and 2 mL of bile was extracted and stored in a sterile cryopreservation tube in liquid nitrogen at -80°C.

##### 2.1.3 Gallbladder mucosa

after surgical resection, the gallbladder mucosa was washed with normal saline. The gallbladder mucosa (about 1cm x 1cm in size) was taken and stored in a sterile cryopreservation tube of -80°C liquid nitrogen.

##### 2.1.4 Feces

a disposable sterile stool tube for bacterial community detection was used 1 day before surgery to avoid contact with urine and avoid collecting samples exposed to air for more than 10 minutes. 5 g of fresh feces were collected and immediately stored in -80°C liquid nitrogen.

#### 2.2 Control group

A disposable sterile feces tube for bacterial community detection was used to avoid contact with urine and avoid collecting samples exposed to air for more than 10 minutes. 5g of fresh feces were collected and immediately stored in liquid nitrogen at -80°C.

Dry ice was used in the transfer process, and the sample environment was kept at -80°C. All experimental personnel involved in specimen collection and transportation received professional training.

### 3. DNA extraction and 16S rRNA amplicon sequencing

DNA was extracted from samples (0.5g) using the QIAamp PowerFecal DNA Kit (QIAGEN, Germany) according to the manufacturer’s protocols. Subsequently, the V3 and V4 region of the bacterial 16S rDNA gene was amplified by PCR using primers xxxx. PCR reactions were performed in a 25 μL mixture containing 5 μL of 5×GC Buffer, 0.5 μL of KAPA dNTP Mix, 0.5 μL of KAPA HiFi HotStart DNA Polymerase, 0.5 μL of each primer (10 pM) and 50∼100 ng of template DNA. PCR reaction cycling included 95 °C for 3 min, followed by 25 cycles at 95 °C for 30 s, 55 °C for 30 s, and 72 °C for 30 s and a final extension at 72 °C for 5 min. AMPure XP beads were used to purify the 16S V3 and V4 amplicon away from free primers and primer-dimer species. The purified product was amplified by PCR using primers xxxx, where the barcode is an eight-base sequence unique to each sample. PCR reactions were performed in a 25 μL mixture containing 5 μL of 5×GC Buffer, 0.75 μL of KAPA dNTP Mix, 0.5 μL of KAPA HiFi HotStart DNA Polymerase, 1.5 μL of each primer (10 pM) and 5 μL of purified product. The PCR reaction consisted of an initial denaturation step at 95 °C for 3 min, followed by 8 cycles at 95 °C for 30 s, 55 °C for 30 s, and 72 °C for 30 s and a final extension at 72 °C for 5 min. The amplicons were subsequently purified by AMPure XP beads to clean up the final library before quantification. Finally, purified amplicons were pooled in equimolar and paired-end sequenced (2×250) on an Illumina MiSeq platform according to the standard protocols.

### 4. Bioinformatics and statistical data analyses

Fast Length Adjustment of SHort reads (FLASH) was used to merge paired-end reads from next-generation sequencing^[13]^. Low-quality reads were filtered by fastq_quality_filter (-p 90 -q 25 -Q33) in FASTX Toolkit 0.0.14, and chimera reads were removed by USEARCH 64 bit v8.0.1517. The number of reads for each sample was normalized based on the smallest size of samples by random subtraction. OTUs were aligned by the UCLUST algorithm with a 97% identity and taxonomically classified using the SILVA 16S rRNA database v128. Alpha and beta diversities were generated in the Quantitative Insights Into Microbial Ecology (QIIME) and calculated based on weighted and unweighted Unifrac distance matrices^[14]^. The linear discriminant analysis (LDA) effect size (LEfSe) method was used to identify species that exhibited statistically significant differential abundances among groups ^[15]^.

### Statistical analysis

SPSS 22 software, Graphpad PRISm7 software and QIIME software were used for statistical analysis, and the Chi-square test was used for categorical data. Two independent samples t-test was used for measurement data after assessing normal distribution and homogeneity of variance between the two groups; otherwise, the Wilcoxon rank-sum test was used. During the multi-group comparison, normally distributed data with homogeneity of variance were analyzed by one-way analysis of variance (ANOVA); otherwise, Kruskal-Wallis rank-sum test was used. Statistical significance was set at P < 0.05.

## Results

### 1. Clinical characteristic

The age, BMI and indirect bilirubin (IBil) of gallstone group were higher than control group (P < 0.05). With the increase of age, BMI and IBil increased, the possibility of gallstone formation increased. There were no significant differences in gender and cholesterol between the gallstone group and the control group.

### 2. OTU dilution curve

The sample dilution curve is a well-established method of random sampling. When certain sequencing quantities are randomly selected, the corresponding diversity index of these sequencing quantities is calculated, and the sequence number and diversity index are used to construct the curve, which is mainly used to compare the richness of sample species under different sequencing quantities. As shown in Fig. 1, when the ordinate (sequencing amount) increased, the ordinate (diversity index) did not increase significantly, indicating that the sequencing amount was sufficient to reflect the overall community structure. Further the sequencing amount did not provide new OTUs.

**Fig.1.**
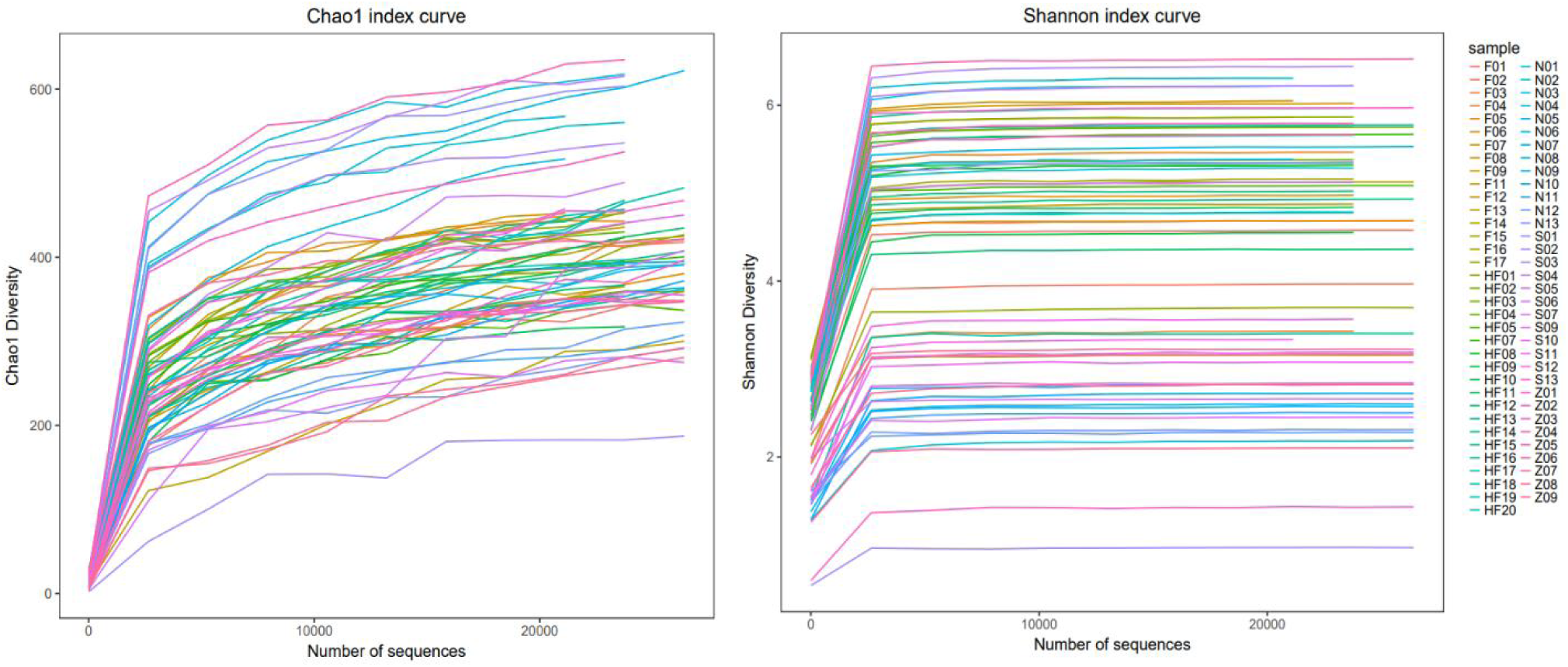
OTU dilution curve

### 3. Bacterial community sequencing results

As shown in Table 2, sequencing of V3-V4 fragments of the 16srRNA gene yielded a total number of sequences of 10 429 883 with a mean ± standard deviation of 151 158±57 813. Among them, 1 818 953 bacterial gene sequences were obtained from gallstones (GSS group), 1 324 611 from bile (GSZ group), 2 089 573 from gallbladder mucosa (GSN group) and 2 163 367 from feces (GSF group). 3 033 379 bacterial gene sequences were obtained from the feces of the gallstone-free control group (HF group).

**Table 1.** Clinical data in gallstone group and control group

| Groups | Gender | Age<br>(years) | BMI<br>(kg/cm<sup>2</sup>) | Cholesterol<br>(mmol/L) | IBil<br>(mmol/L) |
| --- | --- | --- | --- | --- | --- |
| Gallstone group | 8/13 | 52.8±14.4 | 24.4±2.4 | 1.6±0.7 | 8.2±2.2 |
| Control group | 11/9 | 40.1±12.4 | 22.8±2.1 | 1.3±0.5 | 6.9±2.4 |
| P vaule | 0.278 | 0.004 | 0.032 | 0.126 | 0.032 |

**Table 2.**
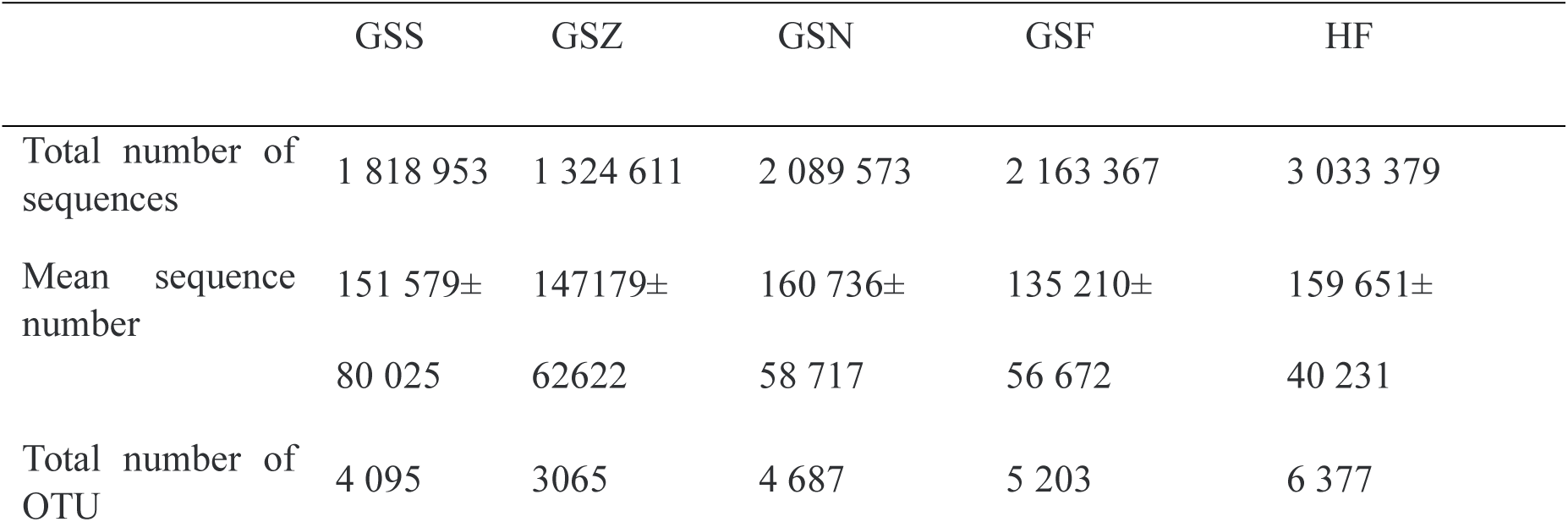
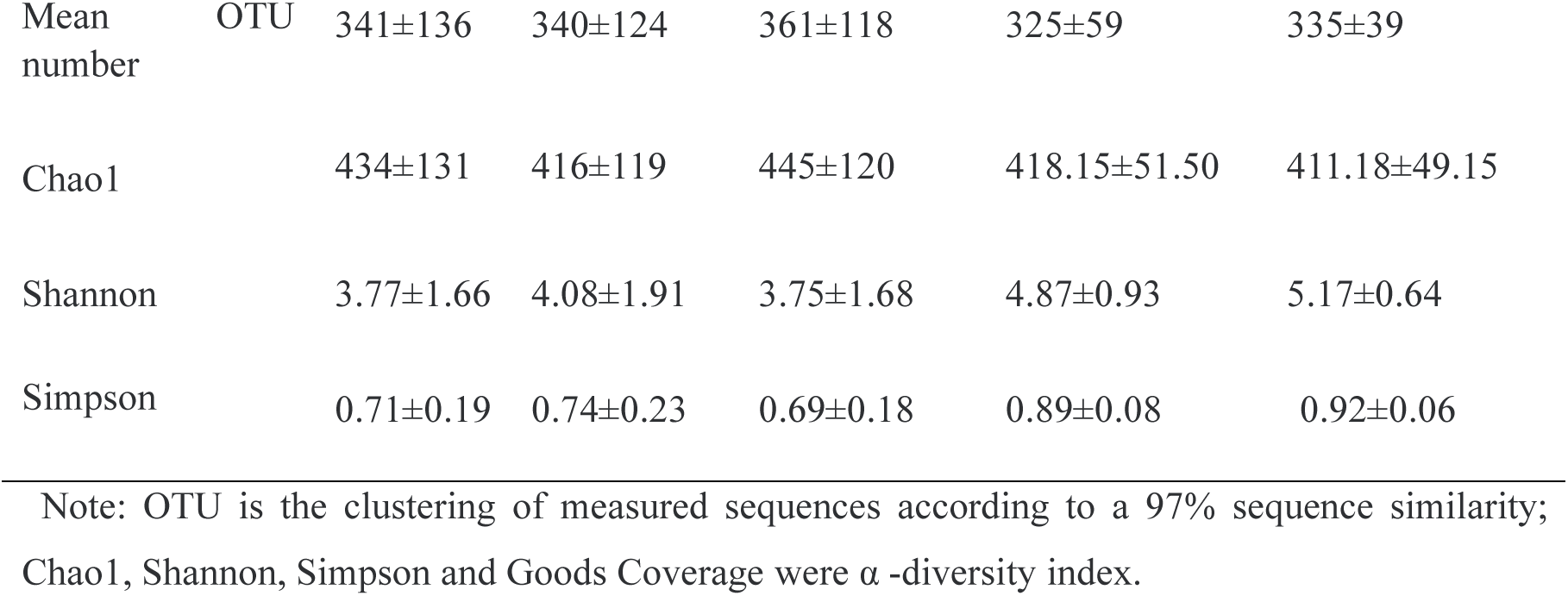
Summary of microflora sequencing results of each group

**Table 3.**
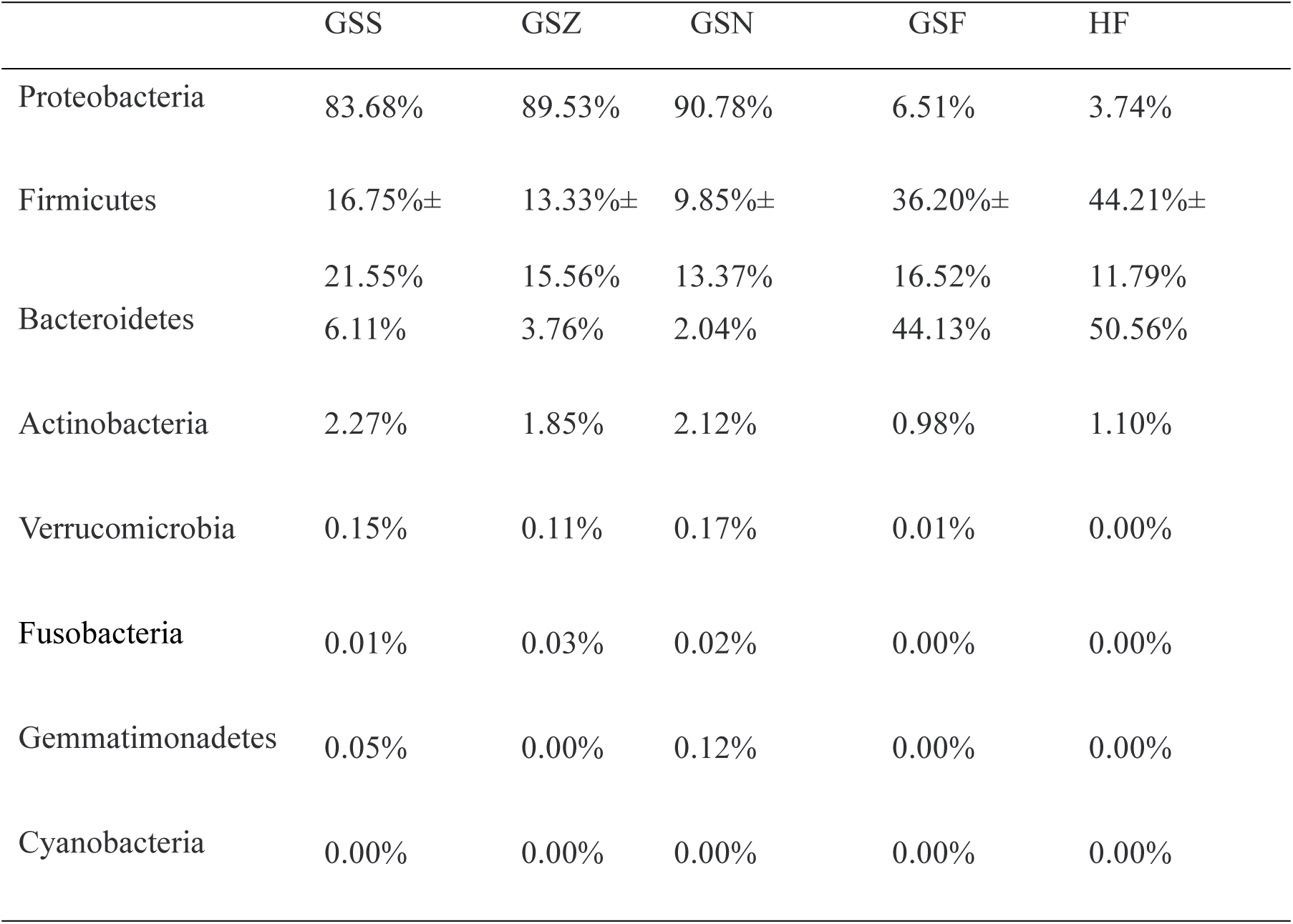
Flora structure among groups (phylum level)

**Table 4.**
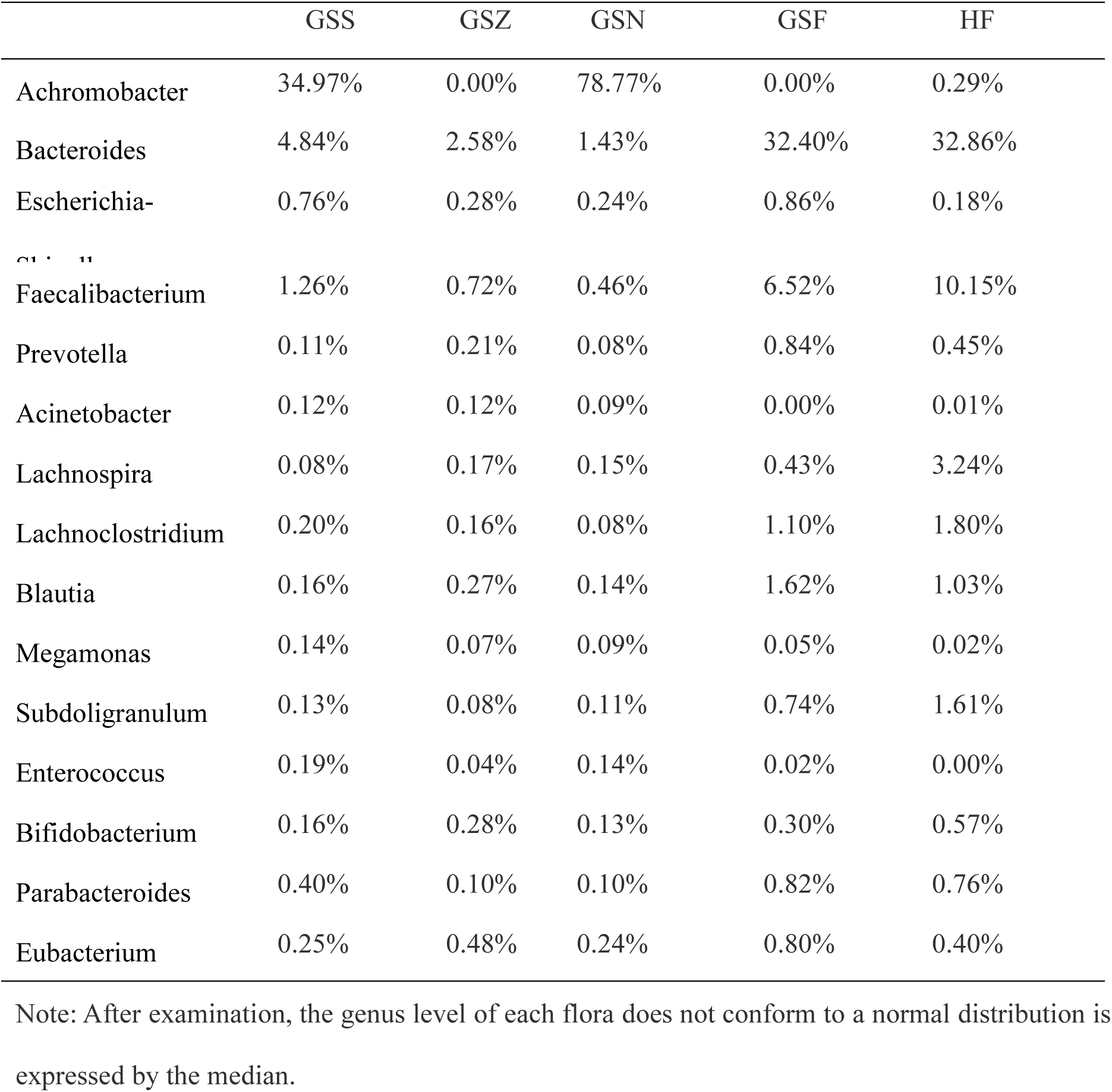
Flora structure among groups (genus level)

### 4. Comparison of intestinal microbiota diversity in patients with gallstones (GSF group) and the control group without gallstones (HF group)

The total sequence number, OTU number, Chao1 index, Shannon index and Simpson index (P > 0.05) showed no significant difference in intestinal microflora diversity between GSF group and HF group.

### 5. Pairwise comparison of bacterial diversity among gallstones (GSS group), bile (GSZ group), gallbladder mucosa (GSN group) and feces (GSF group)

5.1 There was no statistical difference in the bacterial diversity among gallstones, bile and gallbladder mucosa in patients with gallstones.

5.2 There were significant differences in the diversity of gallstones (P=0.004), bile (P=0.045), gallbladder mucosa (P=0.008) and intestinal microflora (Simpson index) in patients with gallstones, and the diversity of intestinal microflora was significantly higher than in the biliary tract (P < 0.05).

### 6. The structure of each group

Biologists classify biology by Domain, Kingdom, Phylum, Class, Order, Family, Genus, and Species. Among these, the highest bacterial taxon is phylum, and the level of phylum in different parts is relatively conservative, which can reflect the difference of flora in different parts. The composition and diversity of bacteria at the genus level are often used to reflect changes in the microenvironment of specific human body parts ^[16]^. Accordingly, we chose to identify the structural analysis of each group of bacteria at the phylum and genus level.

6.1 Comparison of intestinal flora structure at the phylum level between patients with gallstones (GSS group) and the control group (HF group)

Compared with the HF group, Proteobacteria, Firmicutes, Bacteroidetes, Actinobacteria, Verrucomicrobia, Fusobacteria, Gemmatimonadetes and Cyanobacteria (Ps > 0.05) showed no statistical difference in intestinal microflora diversity between patients with gallstones and control group.

6.2 Pairwise comparison of bacterial flora structure at the phylum level in gallstones (GSS group), bile (GSZ group), gallbladder mucosa (GSN group) and feces of patients with gallstones (GSF group)

6.2.1 There was no statistical difference in the bacterial community structure at the phylum level in patients with gallstones, bile and gallbladder mucosa.

6.2.2 As shown in Fig. 2, the abundance of Proteobacteria in the biliary tract was significantly higher, while the abundance of Firmicutes and Bacteroidetes were significantly lower than in the intestinal tract. There was no statistical difference in other microflora structures between the biliary tract and intestinal tract in patients with gallstones.

**Fig.2.**
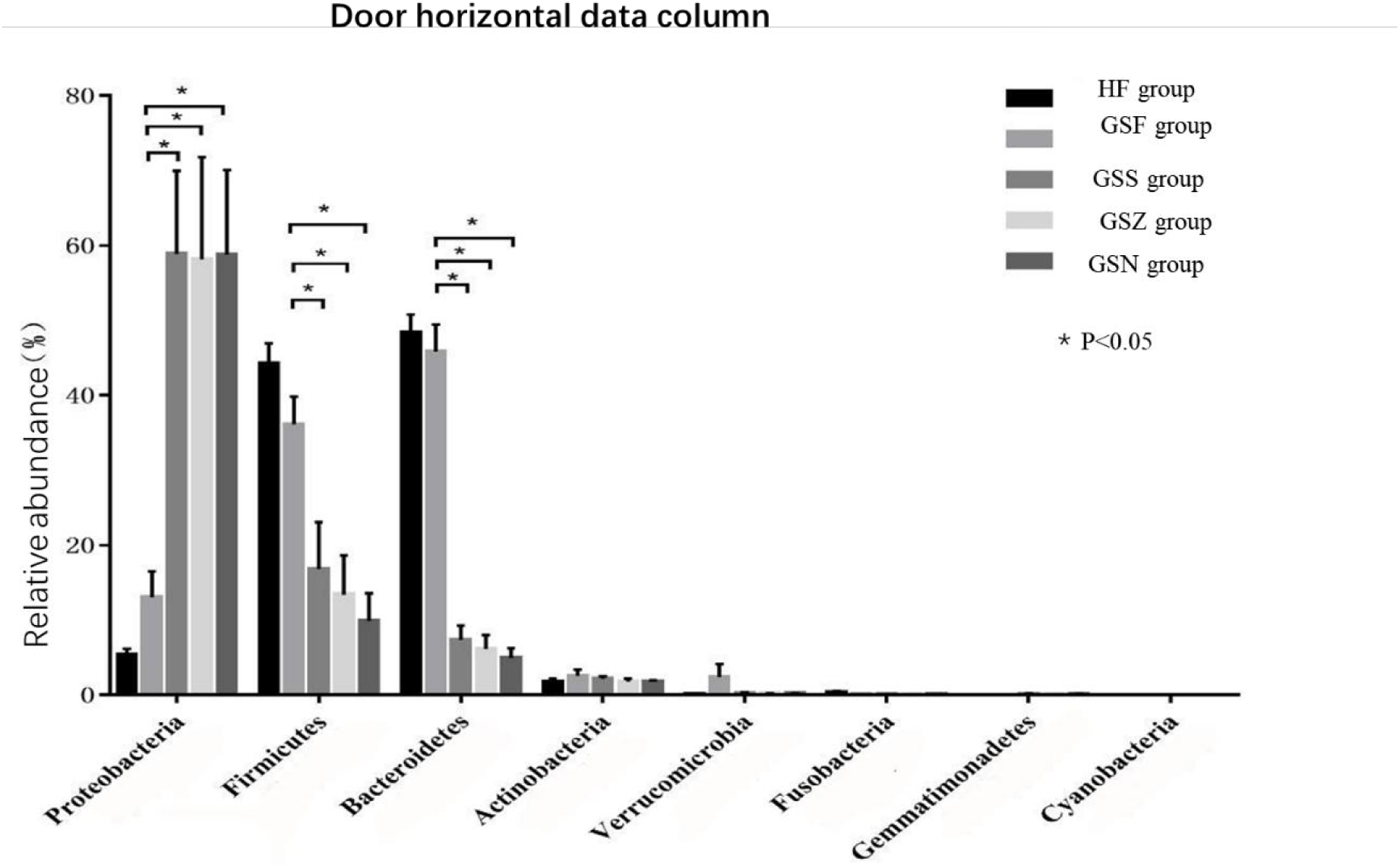
Relative abundance of phyla levels between different groups

**Fig.3.**
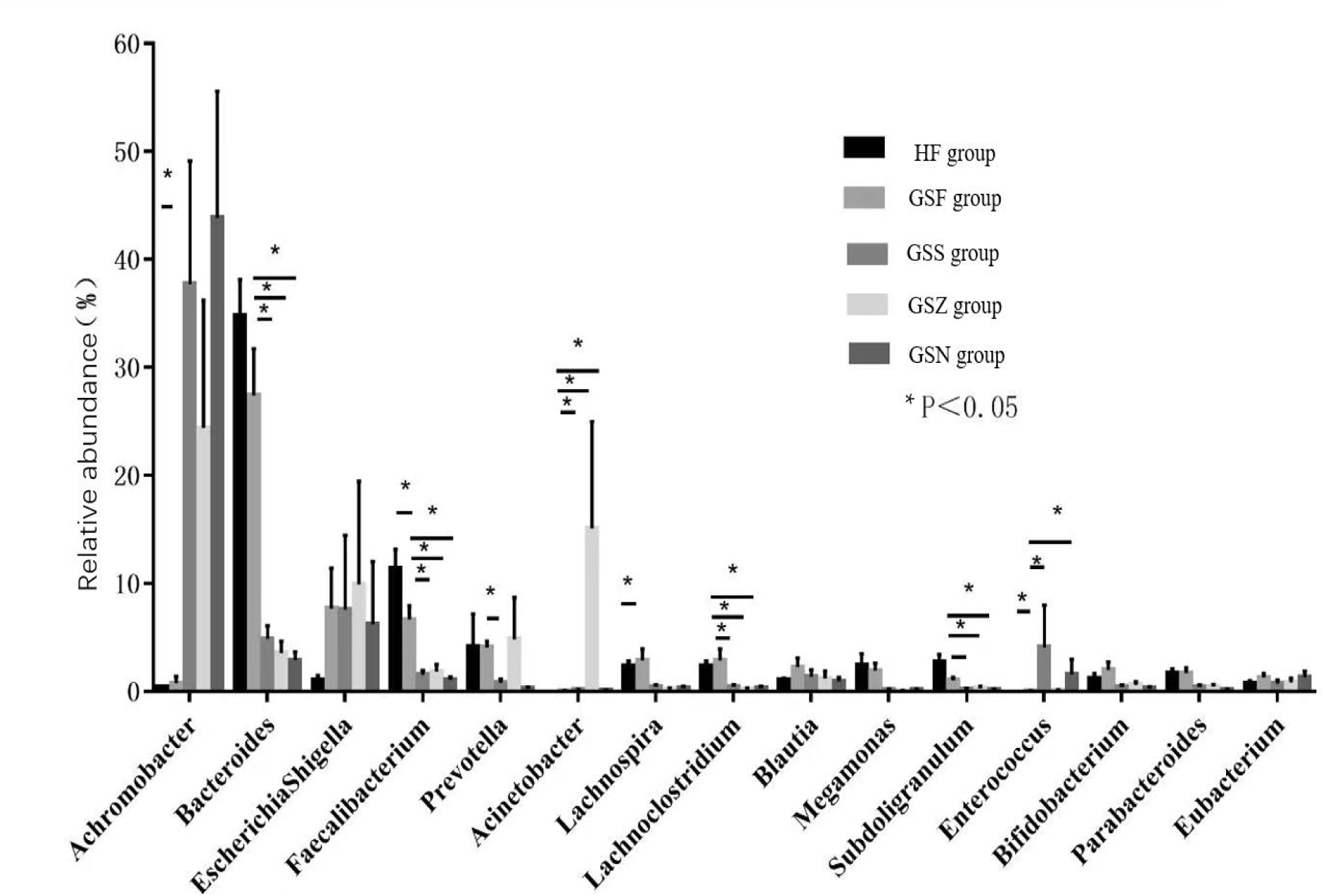
Relative abundance of flora at genus level among different groups

6.3 Comparison of intestinal flora structure at genus level between patients with gallstones (GSF group) and the control group

Compared with the control group, the abundance of Achromobacter, Faecalibacterium and Lachnospira in patients with gallstones was significantly decreased (P < 0.05), and Enterococcus was increased (P < 0.05), while other genera showed no statistical difference(P >0.05).

6.4 Comparison of bacterial flora structure at genus level in gallstone (GSS group), bile (GSZ group), gallbladder mucosa (GSN group) and stool (GSF group) of gallstone patients

6.4.1 Pairwise comparison of gallstones, bile and gallbladder mucosa in patients with gallstones showed no statistical difference in the bacterial community structure at the genus level.

6.4.2 Pairway comparison of calculi, bile, gallbladder mucosa and feces in patients with gallstones:

The abundance of Acinetobacter in the biliary tract (including gallstones, bile and gallbladder mucosa) of patients with gallstones was significantly higher, while Bacteroides, Faecalibacterium, Lachnoclostridium and Subdoligranulum were significantly lower than in the intestinal tract.

In gallstone and gallbladder mucosa of patients with gallstones, the abundance of Enterococcus was significantly higher, but Parabacteroides were significantly lower than in the intestinal tract. However, there was no statistical difference between bile and intestinal flora.

Prevotella in the gallbladder mucosa of patients with gallstones was significantly lower than in the intestinal flora.

### 7. Beta diversity analysis

The above findings suggest that the biliary tract and intestinal flora of patients with gallstones exhibited differences and similarities at both phylum and genus levels. Principal coordinate analysis (PcoA analysis) was conducted based on the Bray-Curtis algorithm to validate these findings. The closer the distance on the axis, the closer the species are.

As shown in Fig. 4, from the PC1 vs. PC2 dimension, the intestinal flora (GSF group and HF group) of patients with gallstones and the control group without gallstones were close. Similar findings were found for the biliary flora (among the GSS group, GSZ group and GSN group). However, the PC2 vs. PC3 dimension showed that the distance between some intestinal flora and biliary tract flora was relatively close.

**Fig. 4.**
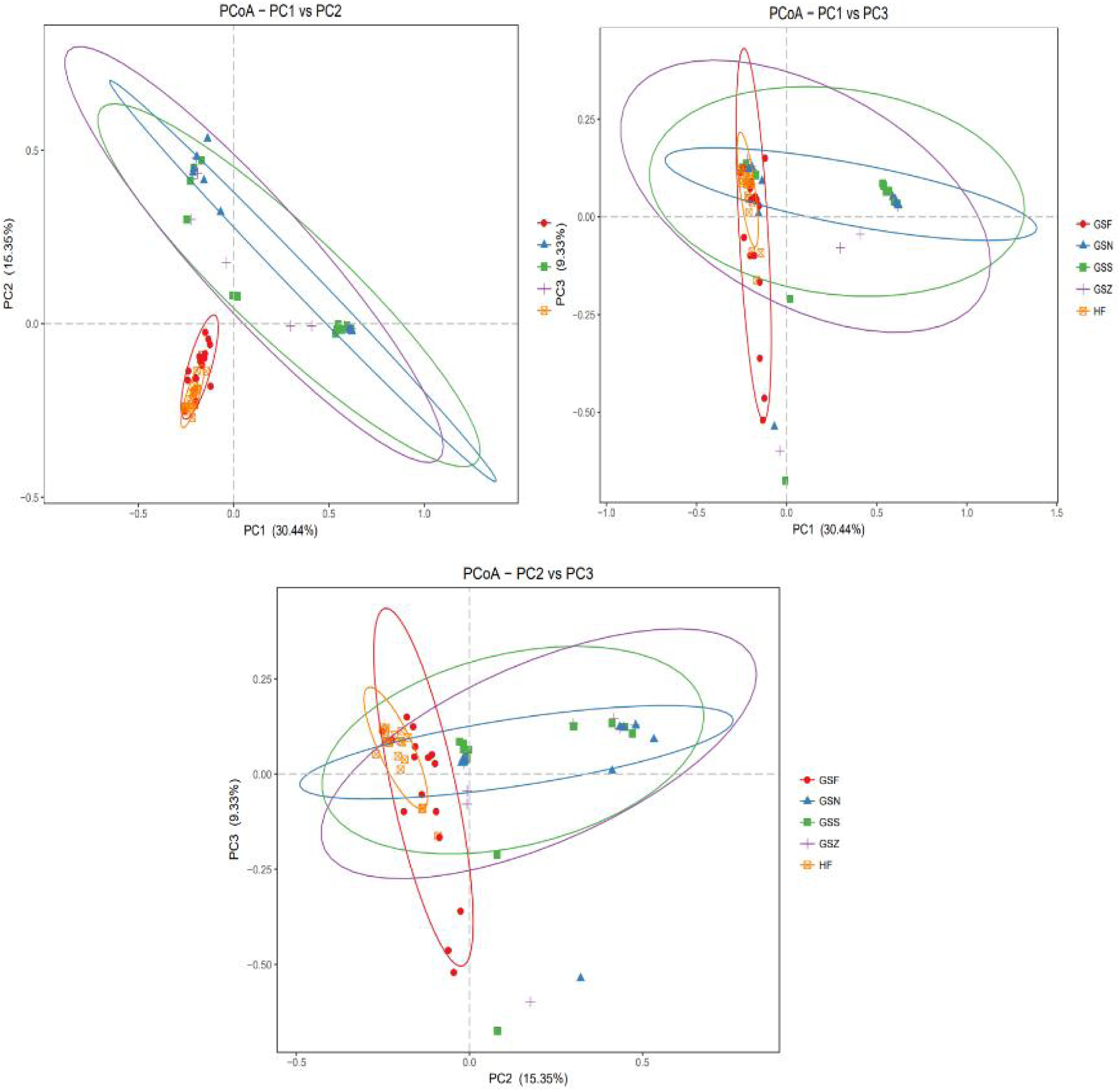
PcoA analysis based on Bray-Curtis algorithm

### 8. Venn plots

OTU with a similarity level of 97% was selected to show the number of OTU shared by multiple groups and reflect the similarity and overlap of environmental samples (Fig. 5).

**Fig. 5.**
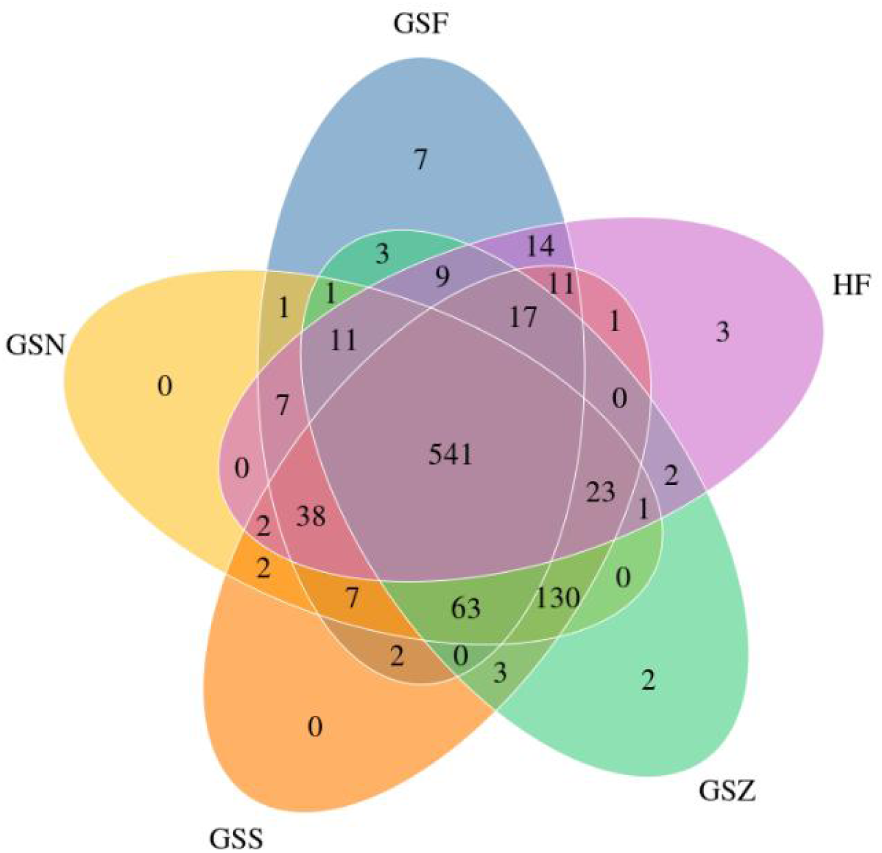
Venn plot:OTUs

The bile and gallbladder mucosa shared 757 OTU, accounting for more than 90% (90.1%, 93.9%, 91.6%) of each group. The intestinal flora of patients and the control group shared 607 OTUs, accounting for more than 85% (87.8%, 95.0%) of each group, respectively. The five groups shared 541 OTUs, accounting for more than 60% of each group (65.6%, 64.4%, 67.1%, 78.3%, 84.7%).

### 9. Difference analysis

The linear discriminant analysis (LDA) was used to reduce the data dimensionality and estimate the impact of the abundance of each species on the difference ^[15]^. Species with LDA values greater than the set threshold were regarded as biomarkers with statistical differences. It is generally believed that an LDA value greater than 3 indicates a significant difference. However, given the large variety of bacteria in bile, gallbladder mucosa, feces of patients with gallstones and feces of the control group, an LDA value greater than 4 was adopted as the threshold value for screening characteristic bacteria (Fig. 6).

**Fig. 6.**
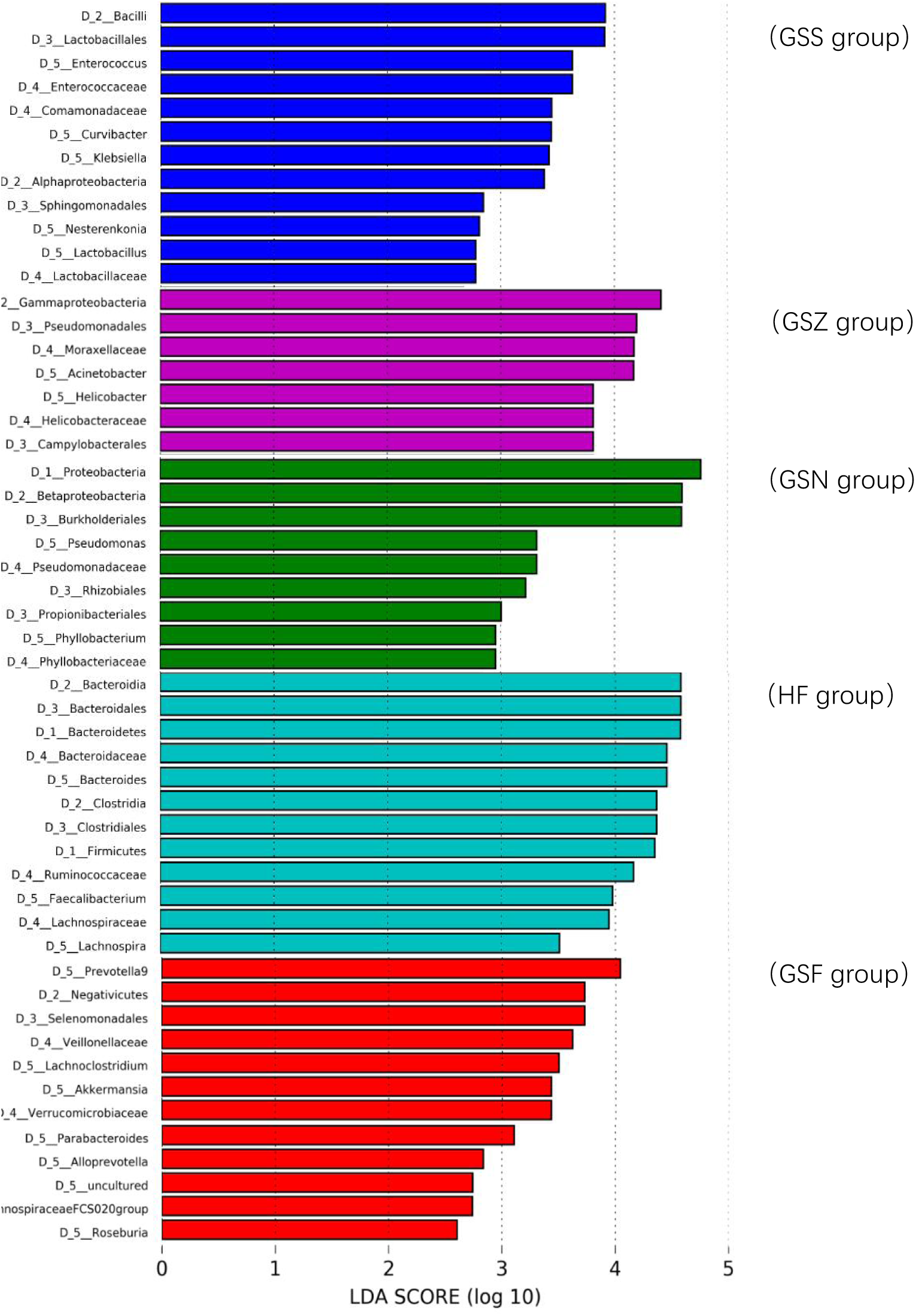
Distribution of LDA scores

1. Prevotella had an LDA value > 4 in the stool group of patients with gallstones.
2. Bacterial species with LDA value > 4 in bile included Gammaproteobacteria, Pseudomonadales, Moraxellaceae and Acinetobacter.
3. Proteobacteria, Betaproteobacteria, and Burkholderiales had LDA > 4 in gallbladder mucosa.
4. No species had an LDA value > 4 in gallstones, while the representative bacteria Bacilli, Lactobacillales, Enterococcus and Enterococcaceae had an LDA > 3.
5. Bacterial species with LDA > 4 in the feces of the control group without gallstones included Bacteroidia, Bacteroidales, Bacteroidaceae, Bacteroides, Clostridia, Clostridiales, Firmicutes, and Ruminococcaceae.

## Discussion

### 1. Risk factors for gallstone formation

Risk factors for gallstone formation include: family history, female, pregnancy, age over 40, etc. ^[17,18]^. Current evidence suggests that people with a family history of gallstones are 5 times more likely to develop gallstones than those without ^[19]^. The incidence of gallstones in women of childbearing age is about twice that of men, and pregnancy puts women at a higher risk of cholelithiasis. About 10% of pregnant women are affected by gallstones^[20]^. However, after menopause, the incidence of gallstones in women gradually approaches that of men ^[19]^. Our study confirmed the conclusion that the gender bewteen patients (pregnant women were not inclued) with and without gallstones had no significant difference. A 4 to 10-fold increased risk of gallbladder disease has been associated with people older than 40 years^[21]^. Obesity, rapid weight loss, high-caloric diet, drugs, type 2 diabetes, metabolic syndrome, dyslipidemia, smoking and sedentary lifestyle have also been documented as influencing factors ^[18]^. Moreover, obesity, especially abdominal obesity, is related to gallstone formation^[19]^.

In this study, we included patients with and without gallstones. No significant differences in gender and cholesterol were found, which were inconsistent with the literature. This discrepancy may be attributed to the relatively small sample size. Moreover, the average age, BMI and IBil of the gallstone group were significantly higher than the control group, consistent with previous research results, given that the elderly are more prone to metabolic diseases such as obesity, type 2 diabetes and dyslipidemia due to reduced activity. Obesity is a major factor in metabolic syndrome and metabolic fatty liver disease, associated with an increased incidence of gallstone formation^[22]^. In a systematic review of patients with gallbladder disease (N= 55,670), the relative risk (RR) for gall bladder-related disease was 1.63(1.49-1.78) for every 5-unit increase in BMI^[23]^. Moreover, the elderly exhibit slower intestinal peristalsis, increasing susceptibility to intestinal flora disorders and may be accompanied by Oddi sphincter relaxation, leading to the retrograde movement of the intestinal flora into the biliary tract.

### 2. Correlation between gallstones and intestinal microflora disorders

Intestinal flora studies have gained momentum in recent years. The human intestine is colonized by 100 trillion bacteria, involved in many body activities. Intestinal dysbiosis has been associated with various human diseases, such as kidney stones, obesity, diabetes, osteoporosis, and polycystic ovary syndrome^[24]^. Most patients with gallstones have no clinical manifestations, but some present with abdominal discomfort such as abdominal distension and belching, which may be related to intestinal flora disorders under the premise of excluding other organic gastrointestinal diseases. The relationship between gallstones and intestinal flora has gradually become a research hotspot.

Given that the intestinal flora is subject to many potential influencing factors, strict inclusion and exclusion criteria were established for this study. For instance, we excluded patients with severe bacteremia, sepsis, and a history of antibiotic or probiotic use in the last 3 months^[25]^. Moreover, we excluded patients with serious comorbidities (metabolic diseases)^[26]^, prior use of somatostatin and other drugs affecting gallstone formation^[27]^, history of intestinal surgery^[28]^ and pregnant women or long-term contraceptive users^[20]^. From the initial cultivation of intestinal and biliary flora to the high-throughput sequencing at present, the methods of bacterial community detection are also changing constantly. In this study, high-throughput sequencing was used to sequence v3-V4 fragments of the 16S rRNA gene. The total number of sequences obtained was 10 429 883, with a mean ± standard deviation of 151 158±57 813. 1 818 953 bacterial gene sequences were obtained from gallstones (GSS group), 1 324 611 from bile (GSZ group), 2 089 573 from gallbladder mucosa (GSN group), and 2 163 367 from feces (GSF group). 3 033 379 bacterial gene sequences were obtained from the feces of the gallstone-free control group (HF group). These findings provide important data for subsequent statistical analysis.

This study found no significant difference in the abundance and diversity of intestinal flora between patients with gallstones and the control group, consistent with the literature^[16]^. However, studies have shown that intestinal microbial diversity decreased, and Roseburia decreased ^[29]^. This discrepancy may be due to the small sample size, emphasizing the need to carry out multicenter studies with large samples for further verification to ensure the robustness of our findings.

In terms of bacterial community structure, there was no significant difference in the intestinal flora structure at the phylum level in patients with and without gallstones. However, at the genus level, the abundance of Achromobacter, Faecalibacterium and Lachnospira in the intestinal flora of patients with gallstones significantly decreased (P < 0.05). Enterococcus was significantly increased (P < 0.05), in which Achromobacter belonged to Proteobacteria, Clostridium tender, Helicobacter pilosa and Enterococcus belonged to Firmicutes. Studies have also shown that proteobacteria in the intestinal tract of patients with gallstones suffer from bacterial overgrowth, including a wide range of pathogenic microorganisms, such as Escherichia coli, Salmonella, vibrio and helicobacter ^[16]^. Animal experiments found that cholesterol stones were formed in mice fed with a lithogenic diet, and the intestinal flora of Firmicutes/Bacteroidetes was significantly reduced ^[30]^. Taken together, the above findings suggest that intestinal flora disorders are common in patients with gallstones.

### 3. Correlation between intestinal flora and biliary tract flora

In this study, the bacterial diversities of gallstones (P=0.004), bile (P=0.045), and gallbladder mucosa (P=0.008) were significantly different from the intestinal flora (Simpson index) (P < 0.05), indicating greater intestinal flora diversity in patients with gallstones was in the biliary tract flora. Some studies have reached a consistent conclusion that the average biodiversity of bile microorganisms in patients with recurrent choledocholithiasis decreases^[31]^, suggesting that the decreased biodiversity may weaken the elasticity of natural ecosystems and increase the possibility of serious ecosystem degradation ^[32]^. Nonetheless, it has been observed that the bacterial diversity of the biliary tract was significantly higher than the intestinal tract^[16]^. This discrepancy may be caused by the significant differences in biliary flora between individuals ^[33]^.

In terms of bacterial community structure, this study found that at the phylum level, the abundance of Proteobacteria in the biliary tract was significantly higher, while the abundance of Firmicutes and Bacteroidetes was significantly lower than in the intestinal tract.The increase of proteobacteria can participate in oxidative stress and is a potential microbial diagnostic marker of epithelial dysfunction ^[34]^. The formation of gallstones is also related to epithelial dysfunction^[35]^, which can explain the high abundance of proteobacteria in patients with gallstones to a certain extent. Growing evidence corroborates that the number of proteobacteria in bile from patients with recurrent choledocholithiasis is significantly higher than in patients without cholelithiasis ^[36]^.

Herein, LefSe analysis (linear discriminant analysis) was used to estimate the impact of sample abundance on the differential effect of each group and to find biomarkers with statistical differences between groups. In this study, LefSe analysis showed many types of bacteria with LDA > 4 in the biliary tract (as shown in Fig. 6), which belonged to proteobacteria. The Firmicutes and Bacteroidetes had an LDA value > 4 in the intestine of the control group. This finding further validated the differences in bacterial community structure at the phylum level.

At the genus level, the abundance of Acinetobacter (belonging to proteobacteria) and Bacteroides, Faecalibacterium, and Lachnoclostridium in the biliary tract were significantly higher than in the intestinal tract. The abundance of Subdoligranulum was significantly lower than in the gut. There is a rich literature available that reveals that Acinetobacter can produce β -glucuronase ^[37]^, which hydrolyzes bilirubin glucuronic acid and produces free bilirubin, which ultimately combines with free calcium ions to form gallstones. This study found a significant increase in Prevotella in the intestines of patients with gallstones compared to the biliary tract flora. In LefSe analysis, Prevotella showed an LDA value> 4 in feces of patients with gallstones, suggesting that Prevotella can be used as a biomarker for bacterial dysregulation in patients with gallstones. In this regard, a meta-analysis of 1791 patients showed that Prevotella is involved in the formation of atherosclerosis ^[38]^ and other related metabolic factors are closely associated with gallstone formation^[11]^.

In this study, common and unique OTU of the biliary tract and intestinal tract were visualized by a venn plot, and it was found that gallstones, bile and gallbladder mucosa shared 757 OTU, accounting for more than 90% of each group. The intestinal flora of the patients and the control group shared 607 OTUs, accounting for more than 85% of each group. The five groups shared 541 OTUs, accounting for more than 60% of each group. The results reflected the similarity and overlap among the samples, indicating that the biliary tract flora may be partly derived from a retrograde intestinal infection, providing novel insights into the source of biliary tract flora. PCoA analysis in this study also fully confirmed this point. As shown in Fig. 5, the PC1 vs. PC2 plots showed that the intestinal flora (GSF group and HF group) of patients and the control group were close, and the biliary tract flora (GSS group, GSZ group and GSN group) was close, suggesting that the biliary tract and intestinal flora exhibited specificity. However, the PC2 vs. PC3 plot showed that the distance between some intestinal flora and biliary tract flora was close, indicating a certain degree of overlap between the intestinal flora and biliary tract flora, providing a foothold for further studies on the source of biliary tract flora.

However, no statistical difference was found in this study on Escherichia coli, which has been documented to be closely related to gallstone formation^[31]^. This discrepancy may be accounted for by the fact that most patients with recurrent common bile duct stones in previous studies underwent ERCP at least once, resulting in the relaxation of Oddi sphincter and the invasion of intestinal microbiota into the biliary tract. Indeed, in the present study, the gallstone patients included underwent gastrointestinal surgery for the first time. Besides, duodenal-bile duct reflux has been associated with the recurrence of common bile duct stones after prior ERCP treatment^[39]^. Interestingly, it was found that bacterial infection in the bile from patients with gallstones accompanied by Oddi sphincter relaxation was more serious and lithogenic, and the bile microbiota could be detected in the upper digestive tract^[40]^.

## Conclusion

The intestinal flora of patients with gallstones and with gallstones exhibited significant imbalance at the genus level. Compared with the intestinal flora of patients with gallstones, the diversity of biliary tract flora was higher. Statistically significant differences in the flora structure were observed at the phylum and genus levels. The biliary tract (gallstones, bile, mucosa) and intestinal flora of patients with gallstones exhibited similarities and differences, providing novel insights into the source of the biliary tract flora. The characteristic bacterial groups in the bile of patients with gallstones were Gammaproteobacteria, Pseudomonadales, Moraxellaceae and Acinetobacter. Proteobacteria, Betaproteobacteria and Burkholderiales were the characteristic bacteria in the gallbladder mucosa of patients with gallstones. The characteristic flora in the feces group of patients with gallstones was Prevotella. The characteristic flora in the feces of the control group consisted of Bacteroidia, Bacteroidales, Bacteroidaceae, Bacteroides, Clostridia, Clostridiales, Firmicutes and Ruminococcaceae. The identification of characteristic flora provides the foothold for further studies on gene function annotation and metabolic pathways to explore the pathogenesis of gallstones and and has certain guiding significance for disease prevention and treatment.

## Data Availability

All data produced in the present study are available upon reasonable request to the authors

## Notes

### Competing Interest Statement

The authors have declared no competing interest.

### Funding Statement

This study was funded by Digestive Medical Coordinated Development Center of Beijing Hospitals Authority (No. XXT14).

### Author Declarations

Clinical research design protocols;the institutional research ethics committee of Beijing Tiantan Hospital, Capital Medical University approval for this work.

